# Novel variants in *ryanodine receptor type 3* predispose to acute rhabdomyolysis due to impaired autophagy

**DOI:** 10.64898/2026.02.27.26345848

**Authors:** Hortense de Calbiac, Laure Caccavelli, Solène Renault, Marine Madrange, Quentin Raas, Marjolène Straube, Guy Brochier, Emmanuelle Lacène, Anaïs Chanut, Angéline Madelaine, Clémence Labasse, Lylia Mekzine, Sebastian Montealegre, Nicolas Goudin, Aleksandra Nadaj-Pakleza, Christel Tran, Stéphanie Gobin, Arnaud Hubas, Apolline Imbard, Pascal Laforêt, Nicolas Dupont, Anne-Sophie Armand, Franck Oury, Filip van Petegem, Teresinha Evangelista, Pascale de Lonlay

## Abstract

Rhabdomyolysis is the acute breakdown of skeletal muscle resulting from failure of cellular homeostasis in response to metabolic stress. Recurrent forms are frequently linked to inherited defects affecting energy metabolism or calcium handling. Ryanodine receptor type 3 (RyR3) is an intracellular calcium release channel, expressed in skeletal muscle, that contributes to the fine-tuning of calcium signaling. Although variants in other calcium-handling proteins have been implicated in rhabdomyolysis, the role of RyR3 has not been established.

In this study, we report rare compound heterozygous missense variants in *RYR3* identified in two unrelated individuals with severe, fever-triggered recurrent rhabdomyolysis. Muscle biopsies revealed mild structural changes with triadic disorganization, mitochondrial alterations, lipid accumulation, and autophagic material, while overall muscle architecture was largely preserved. Structural modeling supports the pathogenicity of the variants, and calcium flux analysis demonstrated significantly reduced ryanodine receptor-mediated calcium release in patient-derived myoblasts.

Functional analyses showed that RyR3 deficiency impaired starvation-induced autophagy, characterized by defective autophagosome formation and reduced autophagic flux, and increased susceptibility to metabolic stress. Mitochondrial bioenergetic profiling revealed reduced oxidative phosphorylation capacity and decreased membrane potential under stress conditions, consistent with compromised mitochondrial adaptation. In zebrafish, *ryr3* knockdown resulted in structural and functional muscle abnormalities, including reduced myotome area and decreased locomotor activity, associated with impaired autophagic flux.

This study establishes a novel association between recessive *RYR3* variants and recurrent rhabdomyolysis and identifies RyR3 as a critical regulator of skeletal muscle stress adaptation through calcium-dependent control of autophagy and mitochondrial homeostasis. More broadly, our findings further highlight autophagy as a central determinant of muscle resilience in the context of rhabdomyolysis.

## Introduction

Rhabdomyolysis (RM) is the acute breakdown of skeletal myofibers in response to a triggering factor, most commonly overexertion, fever, infection, fasting, or certain drugs (notably lipid-lowering agents such as statins)^1^. Typical clinical features include muscle weakness, myalgia, and myoglobinuria. The destruction of skeletal muscle cells dramatically increases serum creatinine kinase (CK) levels and the release of myofiber contents into the circulation^2^. The exact incidence of RM remains difficult to determine due to the lack of prospective studies and likely underrecognition of milder cases.

Importantly, genetic predisposition can underlie acute RM episodes, particularly in pediatric patients with recurrent events^3^. However, the number of known genetic causes remains limited and most cases still lack a molecular diagnosis^3^. Genetic RMs belong to a broader and expanding group of diseases classified as cell trafficking disorders^1^ encompassing defects in intracellular transport, interorganelles communication, and autophagy^1,4^. In these patients, life-threatening RM arises upon metabolic decompensation in response to a stress factor such as fasting or fever. They exhibit normal muscle function between RM episodes, suggesting that baseline muscle metabolism is sufficient to meet cellular demands under resting conditions and that RM occurs when skeletal muscle cells fail to adap to metabolic stress^1^.

The most common genetic causes include metabolic diseases, including fatty acid oxidation disorders^5^, glycogen storage diseases^6^, as well as variants in genes related to skeletal muscle calcium signaling such as *Ryanodine Receptor Type 1* (*RYR1*)^7^, *ATPase Sarcoplasmic/Endoplasmic Reticulum Calcium Transporting 2* (*ATP2A2*)^8^, *SH3 And Cysteine Rich Domain 3* (*STAC3*)^9^, *Calcium Voltage-Gated Channel Subunit Alpha1 S* (*CACNA1S*)^10^, or lipid metabolism such as the *Lipin-1* (*LPIN1*)^11^ and *Transport and Golgi Organization Protein 2 Homolog* (*TANGO2*)^12^ genes.

Ryanodine receptors (RyRs) are intracellular calcium-release channels located on the endoplasmic reticulum (ER), mediating calcium flux into the cytosol^13^. There are three main RyR isoforms^14^: *RYR1* (MIM 180901) the predominant skeletal muscle isoform, located in the sarcoplasmic reticulum (SR) and mitochondria; *RYR2* (MIM 180902) mainly expressed in cardiac muscle cells and neurons^14^; and *RYR3* (MIM: 180903) expressed in the central nervous system and in skeletal muscle^14^.

Pathogenic variants in *RYR1* are associated with acute exertional, febrile or drug-related RM, anesthesia-related malignant hyperthermia syndrome, as well as congenital myopathies^7,15–21^. *RYR2* variants have been linked to cardiac arrhythmias and cardiomyopathies^22–25^, whereas *RYR3* variants have been implicated in congenital myopathy^26^ and arthrogryposis^27^.

In skeletal muscle, *RYR3* expression is developmentally regulated: it is more abundant during the embryonic and early postnatal stages and decreases in adulthood, when RyR1 becomes the dominant isoform^28–31^ RyR3 is located in triads adjacent to RyR1^31^. The triad is a specialized structural complex of the skeletal muscle fibers formed by the close apposition of a transverse tubule (T-tubule) and two terminal cisternae of the sarcoplasmic reticulum (SR). This structure is essential for excitation-contraction coupling (ECC), the process that links membrane depolarization to calcium release and muscle contraction. Upon neuromuscular transmission, the activation of nicotinic acetylcholine receptors at the motor endplate depolarizes the sarcolemma, which propagates along the T-tubules. This depolarization activates voltage-dependent L-type calcium channels (dihydropyridine receptors, DHPRs), which are mechanically coupled to RyRs on the SR membrane. The activation of RyRs results in the release of calcium from SR stores into the cytosol, initiating muscle contraction^32–34^. In addition to depolarization-driven activation, RyRs are also modulated by intracellular signals and pharmacological agonists, contributing to the regulation of calcium dynamics in skeletal muscle^35–39^. Unlike RyR1, which is mechanically coupled to DHPRs, RyR3 is thought to contribute primarily through calcium-induced calcium release: calcium released through RyR1 can activate nearby RyR3 channels, thereby amplifying and modulating the overall calcium signal. RyR3 is therefore believed to fine-tune calcium release, and influence the spatial and temporal properties of calcium transients^28,33,37,40–42^.

Autophagy is an evolutionarily conserved lysosomal degradation pathway that mediates the turnover of cytoplasmic components, including proteins, protein aggregates, and damaged organelles. Through the formation of double-membrane autophagosomes that subsequently fuse with lysosomes, autophagy enables the recycling of intracellular substrates and supports cellular energy homeostasis, particularly under conditions of nutrient deprivation or metabolic stress. In skeletal muscle, autophagy plays a critical physiological role in maintaining fiber integrity, regulating organelle quality control, and adapting to fluctuations in energetic demand. Autophagy is essential for maintaining muscle homeostasis and adaptation to stress, and defects in this process have been reported in a large spectrum of skeletal muscle disorders, including RM where failure to respond properly to metabolic stress predisposes muscle fibers to breakdown^1,43–48^.

Here, we report two unrelated individuals with recurrent RM harboring rare compound heterozygous variants in *RYR3*. Given the functional proximity of RyR3 to RyR1 within the calcium release unit and its role in shaping intracellular calcium dynamics, we hypothesized that *RYR3* variants impair skeletal muscle stress adaptation and thereby predispose individuals to RM. Knowing that calcium signaling is a critical upstream regulator of autophagic activation and metabolic remodeling in skeletal muscle^49–55^, we postulated that disruption of RyR3-mediated calcium release might compromise autophagic responses to energetic stress, thereby increasing muscle vulnerability.

Accordingly, we show that these variants impair RyR3-mediated calcium release and are associated with triadic structural abnormalities in skeletal muscle. Using patient-derived myoblasts and a zebrafish loss-of-function model, we show that RyR3 deficiency disrupts starvation-induced autophagy, alters mitochondrial bioenergetics, reduces mitochondrial mass and biogenesis signaling, and increases susceptibility to metabolic stress. Together, our findings identify *RYR3* as a novel gene associated with recurrent RM and reveal an unrecognized role for RyR3 in coordinating the calcium-dependent autophagic and mitochondrial pathways essential for skeletal muscle homeostasis.

## Methods

### Study approval

Patient biopsies and myoblasts were obtained during diagnostic procedures and clinical care. Informed consent was obtained from all patients, and donors provided free and informed consent for the storage of biological samples and associated health data. The biobank has been declared to the Ministry of Research (IE-2011-593) and updated under the codecoh registration (INSERM) # DC-2025-7811.

### Patient details and investigations

We clinically characterized two patients who presented with severe episodes of RM beginning in adolescence. We performed pedigree analysis and a neurological examination, including muscle strength evaluation using the Medical Research Council (MRC) grading scale. Serum CK levels were measured during acute episodes and at baseline between episodes. Cardiac investigations and muscle biopsy were also performed to establish a diagnosis.

### Histopathological investigations

Muscle tissue was obtained from a deltoid biopsy as part of routine diagnostic investigations at least two months after the first initial RM episode. The samples were frozen in liquid nitrogen-chilled in isopentane and processed for routine histological and histochemical techniques. Electron microscopy analysis was performed on muscle biopsy specimens that were postfixed with glutaraldehyde (2.5 %, pH 7.4), post-fixed with osmium tetroxide (2%), dehydrated in a graded series of ethanol and embedded in epoxy resin (Epon, Electron Microscopy Sciences, USA). Ultrathin sections (90 nm thick) were collected on nickel grids and stained with uranyl acetate and lead citrate. The grids were observed using a “JEOL 1400 Flash” transmission electron microscope (120 kV), and the sections were photodocumented using a Xarosa digital camera (SoftImaging System, France).

For neutral lipid staining, skeletal muscle sections were fixed in Baker’s fixative (36.4% formaldehyde, calcium chloride, and distilled water) and subsequently processed in 70% ethanol. Oil Red O staining was performed using a solution prepared by dissolving 3 g of Oil Red O in 70% ethanol and incubating it at 37°C prior to use.

### Myoblast culture

Myoblasts were obtained from patient muscle biopsies and from 3 age-matched control individuals. Myoblasts from an additional patient with recurrent exercise-triggered RM (CK 90,000 IU/L in their twenties) and carrying a heterozygous *RYR1* variant were also investigated.

The pathogenic variant (NM_00540.2, exon 17, c.1840C>T (p. Arg614Cys)) is well-characterized in RM and malignant hyperthermia and RM^56^. Human primary myoblasts were isolated and grown as previously described^48^. They were cultured in growth medium (GM) (Ham’s-F10 with GlutaMAX, ref: 41550021, Thermo Fisher Scientific) containing 20% FBS (ref S1300-500: Biowest) or in Earle’s balanced salt solution (EBSS, ref: 24010043, Thermo Fisher Scientific) to induce starvation.

### Genetic analyses

DNA was extracted from the peripheral blood leukocytes of the two affected individuals and their parents after providing informed consent, and whole-exome sequencing was performed as previously described^57^. Bioinformatic analysis was performed to detect both qualitative and quantitative variants. The potential impact of novel variants at the protein level was assessed *in silico* using Polyphen 2, CADD, ClinVar, SpliceSiteFinder-like, MaxEntScan, NNSSPLICE, and GeneSplicer. Putative pathogenic variants in *RYR3* were confirmed by Sanger sequencing.

### Gene expression analysis in myoblasts

Total RNA was isolated from patients and control primary or immortalized myoblasts using NucleoSpin RNA XS (Macherey-Nagel) according to the manufacturer’s instructions. Single-stranded cDNA was synthesized from 1 μg of total RNA using the High-Capacity RNA-to-cDNA Kit (Applied Biosystems) after the depletion genomic DNA. The expression of the *RYR3* gene in myoblasts was assessed via RT-qPCR with the following primers: *RYR3*: Fw 5’ GCCATGCAAGTAAAGTCTGGA G 3’ and Rv 5’ GTATCGCGATTTTGCGAGGG 3’. RT-qPCR was performed with Power SYBR® Green PCR Master Mix, and the results were normalized to the *actin beta* gene, and quantified using the 2^ΔΔct^ method.

### Structural analysis of RYR3 variants

To analyze the variants in the context of the 3D structure of the RyR3, the following PDB files were used: 4ERV (crystal structure of the human RyR3 Repeat12 domain, corresponding to residues 2591-2800)^58^ and 9C1E (cryo-EM structure of Mink RyR3 in the closed conformation and bound to FKBP12.6)^59^. Visualization of the side chains of the sequence variants and their environments, along with calculation of hydrogen bonds and salt bridges were performed using Pymol (Schrodinger). The domains were colored according to the domain assignment for RyR3 as previously described^59^.

### *RYR3* knockdown in immortalized human myoblasts

Immortalized human myoblasts from healthy individuals were cultured as described in Mamchaoui *et al*., 2011^60^. siRNAs targeting the RyR3 gene (ref L-006294-00-0005, Horizon Discovery) or a scramble control siRNA (ref D-001810-01-20, Horizon discovery) were transfected into myoblasts in 6-well plates via DharmaFECT transfection reagent (ref T2001-02, Horizon Discovery). After 3 days, the inhibition of *RYR3* expression was assessed via RT-qPCR (see primer sequences above) and the transfected myoblasts were then seeded in gelatin-coated 96-well plates for differentiation assays (20000 cells/well)

### Myoblast differentiation

Differentiation was induced by replacing growth medium with DMEM supplemented with 10 μg/ml insulin (ref 91077-C, Merck) and 1% penicillin-streptomycin (ref P4333, Merck). NuncLight Red reagent for nuclear labeling (ref 4717, Essen Biosciences Ltd, UK) was added in each individual well at a 1/1,000 final dilution. Fluorescence and phase images were taken using IncuCyte® S3 Life Cell Analysis System (Essen Biosciences Ltd, UK). Differentiation was assessed by the following fusion index (number of nuclei per cell with more than 3 nuclei/total number of nuclei).

### Calcium release assay

The cells seeded on an Ibidi µ-Slide 8 Well (Ibidi, Clinisciences) were stained with 5 μM Fluo-4 AM (Invitrogen^TM^) for 30 min to 1h at 37°C in a 5% CO_2_ incubator following the manufacturer’s instructions. After being washed in PBS, the cells were cultured in Krebs-Ringer’s solution (Krebs-ringer bicarbonate buffer, Sigma Aldrich), and the plates were transferred to a temperature-controlled stage (Harvard Apparatus, Holliston) on a confocal Leica SP8 inverted microscope. Cultures were visualized using a 40X oil objective. The intracellular probes were excited at 488 nm, and the emission signals were detected with a 530 ± 30 nm filter. Fluo-4 signals were recorded for 2 min before stimulation to obtain baseline fluorescence values. Then, RyRs were activated by injection with 4-chloro-m-cresol (4-CmC) (Sigma‒Aldrich)^35^, and the fluorescent signal was recorded for 10 min. Ionomycin (1 µg/mL) and EDTA were used as positive and negative controls, respectively. Cytosolic calcium was expressed as deltaF_0_/F_0_, with F_0_ representing the mean fluorescence level before injection. The peak cytoplasmic calcium release and the area under the curves after injection were calculated. Microsoft Excel software (Microsoft Corporation, Redmond, WA, USA) was used to correct the baseline values of Fluo-4.

### Cell death assay

Myoblasts were cultured in GM or upon starvation (EBSS, 4h) to test for potential cell death. The cells were then detached with trypsin and stained in PBS with 5 µM Caspase 3/7 Green Detection Reagent (Thermo Fisher Scientific), after which iodure propidium was added to label both necrotic and apoptotic cells. The fluorescence was recorded on a BD LSR Fortessa flow cytometer and analyzed with FlowJo software.

### Western blotting

To mimic environmental stress, the GM was replaced with EBSS medium with or without 100 nM bafilomycin A1 (ref: tlrl-bafA1, InvivoGen) for 1h and 3h. Myoblasts were lysed in RIPA buffer supplemented with protease inhibitors as previously described. Whole-cell extracts were quantified by BCA before loading equal amounts of protein were loaded per lane onto NuPage 4-20% Bis-Tris gels (ThermoScientific). Dry transfer onto PVDF membranes was performed with an iBlot2 device according to the manufacturer’s instructions (ThermoScientific). Primary antibodies against LC3B (clone 4E12, MBL International, 1:1000) and β-actin (sc-81178, Santa Cruz Biotechnology, 1:1000) were detected with appropriate horseradish peroxidase (HRP)-conjugated secondary antibodies followed by enhanced chemiluminescence (ECL) detection. When samples could not be resolved on the same gel, the gels were run simultaneously and the corresponding membranes were exposed at the same time to ensure identical detection conditions. To guarantee that the blots were measured in the linear range of exposure, all the blots were quantified at the maximal exposure level below saturation via ImageLab software (Bio-Rad).

### Autophagosome staining

Primary myoblasts were cultured in GM or in EBSS (2h). The cells were fixed and subjected to immunofluorescence with an anti-LC3B antibody (clone 4E12, MBL International, 1:100) as previously described^48^. Images were acquired via confocal fluorescence microscopy on a Spinning Disk system (Intelligent Imaging Innovations, USA), an Examiner.Z1 upright stand (Carl Zeiss, Germany), a CSU-W1 head (Yokogawa, Japan), and an ORCA-Flash 4.0 camera (Hamamatsu, Japan), with a 63x (NA 1.4) oil immersion objective. Positive LC3 vesicular signal was recognize first with machine learning pixel classification with ilastik (v1.3.3 post3) and exported as labelmap in order to perform LC3 vesicule segmentation and measurments within an Icy v1.9.5.1 pipeline. Quantification was performed with Icy software (v1.9.5.1 BioImage Analysis unit, Institut Pasteur, France). A total of 15-20 cells were imaged per condition.

### Mitochondrial function in primary myoblasts

Seahorse measurements were performed as previously described^36^. Changes in mitochondrial membrane potential in myoblasts in basal and stress condition (4h EBSS, 20 min FCCP) were measured using TMRE (Abcam-113852) following the manufacturer’s instructions.

### Maintenance of Zebrafish

Adult and larval zebrafish (*Danio rerio*) were maintained at the Imagine Institute (Paris) facility and bred according to the National and European Guidelines for Animal Welfare. Experiments were performed on wild-type and transgenic zebrafish larvae from AB strains. The zebrafish were raised in embryo medium: 0,6 g/L aquarium salt (Instant Ocean, Blacksburg, VA) in reverse osmosis water containing 0,01 mg/L methylene blue. The experimental procedures were approved by the National and Institutional Ethical Committees. Zebrafish were staged in terms of hours post-fertilization (hpf) based on the basis of morphological criteria and manually dechorionated using fine forceps at 24 hpf. All the experiments were conducted on morphologically normal zebrafish larvae.

### *ryr3* inhibition in zebrafish

A morpholino (MO) antisense oligonucleotide (GeneTools, Philomath, USA) was used to specifically knockdown the expression of the *ryr3* zebrafish orthologue. The sequence is 5’ CTTTTACTGACATACCCACTGCTAC 3’. A control MO with no target sequence in the zebrafish genome has the following sequence 5’ ATTCTCGAGCACAGCCTCAAGACAT 3’. Both MOs were injected at a final concentration of 0,6 mM. Microinjections were carried out at one cell stage. To assess the efficiency of ryr3-MO, total RNA was extracted from samples using standard procedures and reverse-transcribed into cDNA according to the manufacturer’s instructions. PCR amplification was performed via primers flanking the exon–exon junction of interest to assess alternative splicing events. The PCR products were separated via 1% agarose gel electrophoresis (the primer sequences are provided in the following section).

### Gene expression analysis in zebrafish

Total RNA was isolated from the injected fish via TRIzol Reagent (Sigma) according to the manufacturer’s protocol. First-strand cDNAs were obtained by via reverse transcription of 1 μg of total RNA via a high-capacity cDNA reverse transcription kit (Roche), according to the manufacturer’s instructions. The expression of the *ryr3*, *trim63* and *ppargc1a* genes was assessed via RT-qPCR with the following primers: *ryr3*: Fw 5’ CTGCACCTCTCCATCTCCAA 3’, Rv 5’

TACCGGCCTCATAGTTCACC 3’; *trim63*: Fw 5’ CCTGGCTTTGAGAGTATGGACC 3’, Rv 5’ GCCCCTTGCCTCACAGTTAT 3’ and *ppargc1a:* Fw 5’ CAGTTCTGGTGGCAGGAAAC and Rv 5’ GTGGATGTAGTCGCCGAGTA 3’ using Power SYBR® Green PCR Master Mix, normalized to *actin beta* gene, and quantified via the 2^ΔΔct^ method.

### Zebrafish locomotor analysis

The motor behavior of 50 hpf zebrafish larvae was assessed via the Touched-Evoked Escape Response (TEER) test, as previously described. Briefly, zebrafish were touched on the tail with a tip and the escape response was recorded via a Grasshopper 2 camera (Point Gray Research, Canada) at 30 frames per second. Distance was quantified frame per frame for each embryo via the video tracking plugin of FIJI 1.83 software (open source). For drug treatment experiments, 30 hpf zebrafish embryos were raised in embryo medium containing 0,3 μM and 0,6 μM atorvastatin (ATV, Sigma-Aldrich, CAS 134523-03-8) or 50 μM metformin (Sigma-Aldrich, CAS: 1115-70-4) dissolved in DMSO and locomotor phenotype was assessed at 50 hpf as described above. The locomotor phenotype results obtained following ATV and metformin treatments were derived from the same experiments and therefore shared the same control groups.

### Autophagic flux in zebrafish

To monitor autophagy flux in zebrafish, we coinjected the GFP-LC3-RFP-LC3ΔG probe developed by Mizushima’s laboratory^61^ with ryr3 or control-MO at a final concentration of 120 ng/μL. GFP and RFP fluorescent signals were captured from the myotomes of live 48 hpf zebrafish larvae using a spinning disk system (see above), with a 20x objective (NA 1.0). The stacks were processed with FIJI 1.47 software, and the same treatments were applied for all the conditions.

Maximal Z intensity projections were created from stacks with FIJI v1.47. Then, as an indicator of autophagy efficiency, RFP fluorescence intensities were normalized to the corresponding GFP signal by calculating the RFP/GFP ratio using the FIJI calculator plugin v1.83. Measurements were performed in two randomly selected regions of interest within skeletal muscle per embryo, with nine embryos analyzed per condition

### Immunofluorescence and Oil Red O staining of zebrafish sections

50 hpf embryos were anesthetized in 0,2 % tricaine, fixed in 4% PFA and prepared for cryosectioning as previously described^48^. The samples were cut into 20 μm thick transverse sections, which were blocked and permeabilized with 0.2% gelatin and 0.25% Triton X-100 (diluted in 1X PBS and incubated overnight with the following primary antibodies: anti-myosin heavy chain (MHC, 1:25 DSHB MF20) to label skeletal muscles and anti-TOMM20 (1:200, rabbit polyclonal, HPA011562 Sigma Aldrich, USA). After several washes, the sections were incubated for 1h with the appropriate secondary antibodies conjugated to Alexa Fluor® (1:500, Thermo Fisher Scientific). Oil Red O (ORO) staining was performed on zebrafish cryosections to visualize neutral lipid accumulation according to standard procedures. The sections were incubated with freshly prepared Oil Red O working solution, rinsed to remove excess dye, and mounted for imaging. The sections were rinsed and mounted in Fluoromount-G™ medium (Thermo Fisher Scientific). Images of the myotome area were captured with a spinning disk system (see above) with a 63X objective. Standard deviation Z projections images were created from stack images per embryo with FIJI 1.83 software and same treatments were applied for all the conditions. Skeletal muscles were manually delineated based on the MHC signal, and their cross-sectional area was quantified via FIJI. TOMM20 immunofluorescence was segmented via FIJI with identical thresholding parameters applied to the mis-MO and ryr3-MO conditions. The ORO signal (lipid droplets) was analyzed using the same approach. The “Analyze Particles” plugin was subsequently used to quantify TOMM20-positive and lipid droplets elements, determine their number, measure mitochondrial size, and calculate the percentage of skeletal muscle area occupied by the respective signals. Five to sixteen zebrafish larvae per condition were analyzed.

### Statistical analysis

The data were plotted and analyzed with R or with Prism (GraphPad) softwares. All the experiments were conducted at least three times unless otherwise specified.

## Results

### Clinical phenotypes of the probands

Two unrelated male individuals of French origin families were included in the study (**Table 1**). Both were born at term and had uneventful birth histories and early motor and cognitive development. Each patient experienced recurrent RM episodes precipitated by febrile illnesses, including influenza, except for one episode in Patient 1 (P1), which had no apparent trigger. The first episode occurred during adolescence for both patients. The number of acute RM episodes ranged from two in P1 to eight in Patient 2 (P2). Acute symptoms included generalized weakness, inability to walk, myalgia, and dark urine. Examination revealed marked tenderness over the thighs and calf muscles without erythema. Reflexes were preserved, and the central nervous system was typically unaffected during episodes. During acute RM, plasma CK levels increased markedly (peaks 80,000 and 300,000 IU/L, respectively; N <150), accompanied by myoglobinuria. Lactate dehydrogenase and aspartate aminotransferase levels were also elevated. Urea, creatinine, uric acid, plasma lactate, total/free carnitine, the acylcarnitine profile, plasma amino acids, and urinary organic acids were normal (data not shown). Intravenous fluids and urine alkalinization maintained normal creatinine and urinary output.

**Table 1.** Clinical presentation and skeletal muscle ultrastructural alterations associated with *RYR3* variants.

| Patients | P1 | P2 |
| --- | --- | --- |
| *Clinical features* |  |  |
| Sex | M | M |
| Age of onset (years) | 15-20 | 15-20 |
| RM episodes | 2 | 8 |
| Triggers | Infection | Infection |
| Creatine kinase peak (IU/L) | 80 000 | 300 000 |
| Mutations | p.Arg2669His and p.Asn3849His | p.Val610Ile and p.Val2986Ile |
| *Ultrastructural features* |  |  |
| Sarcolemma | Preserved | Preserved |
| Sarcomeric structure | Preserved | Relatively preserved. One fiber showed sarcomere misalignment with focal mitochondrial loss. One fiber showed Z-line smearing. |
| Fiber size variability | Yes | Yes, mild |
| Atrophic fiber | Yes, isolated | Yes, isolated |
| Internalized nuclei | No | No |
| Predominance of a fiber type | No | No |
| Triads | Occasional disoriented triads, with sparse dilatation and rare areas of increased density | Dilated sarcoplasmic reticulum (>200 nm), rare triad duplications |
| Autophagy | Mitophagy-like structures, autophagic debris | Autophagy vacuoles, mitophagy-like structures |
| Mitochondrial features | Subsarcolemmal accumulation, some vacuolated mitochondria, presence of myelinoid bodies | Vacuolated mitochondria, presence of myelinoid bodies |
| Myelinoid bodies | Yes | Yes |
| Filamentous body | - | Yes (one fiber) |
| Lipid content | Slightly increased, scattered lipid droplets | Slightly increased, scattered lipid droplets |
| Free glycogen | Yes | Yes |

Between episodes the patients were in good health. Weight, height and intellectual development were normal. Physical exams, including muscle strength tests, were normal; they took part in physical activities. The plasma CK levels were within or near normal (P1: 121-284 IU/L; P2: 200-300 IU/L; N<160 IU/L). P2 reported mild fatigability with occasional myalgia after intense exertion. Interictal EMG, echocardiogram, pulmonary function tests, and (for P2) muscle MRI were normal.

### Molecular investigations

To identify the causative gene mutation, we first excluded fatty acid oxidation defects by biochemical investigation. We then screened a RM gene panel of 51 genes associated with metabolic and calcium-related RMs (including *RYR1, CACNA1S*, and *STAC3)*, selected from the literature and other databases (OMIM, NCBI Gene, Pubmed, HGMD, Genetics Home Reference), via next generation sequencing (NGS, Illumina HiSeq with Agilent probes); no pathogenic variants were found. Exome sequencing of P1 and P2 revealed *RYR3* (NM_001144869.2) as the sole candidate gene, by its function. In P1, three heterozygous variants were found: c.8006G>A (p. Arg2669His), c.11545A>C (p. Asn3849His), and c.10707-102A>T. The c.8006G>A variant is predicted to be deleterious, rare in controls (0,0012%), and inherited from his mother in cis with the rare intronic c.10707-102A>T variant, for which splicing algorithms predict no major effect (SpliceSiteFinder-like; MaxEntScan; NNSSPLICE; GeneSplicer). The c.11545A>C variant is predicted to be benign (CADD, PolyPhen2, ClinVar), is relatively frequent in controls (0,28%, gnomAD), and was inherited from his father. In P2, two heterozygous variants were identified: c.1828G>A (p.Val610Ile) and c.8956G>A (p.Val2986Ile). Both are predicted to be damaging by *in silico* tools (CADD, PolyPhen-2) and are relatively frequent in the general population (∼0.22%). c.1828G>A was inherited from the mother and c.8956G>A was inherited from the father. Both variants were confirmed by bidirectional Sanger sequencing. Familial testing revealed that the healthy parents and siblings each carry only one of the variants. None of these variants were observed in the homozygous state in >6,000 exomes sequenced at the Imagine Institute (Paris, France).

### Ultrastructural analysis of probands skeletal muscle reveals RM features

Muscle biopsies were performed at distance from the RM episode (>2 months). Ultrastructural analysis of skeletal muscle biopsies by electron microscopy revealed consistent abnormalities despite overall preserved sarcolemmal integrity and largely maintained sarcomeric organization (**Table 1**, **Figure 1**). Mild fiber size variability and isolated atrophic fibers were observed in both patients but the most predominant features were enlarged and disorganized triads in P1 and P2 (**Table 1**, **Figure 1**). Triad abnormalities specifically involve the calcium release unit, supporting the pathogenic relevance of *RYR3* variants as they mirror the spectrum of triadic defects described in *ryanodine receptor* 1 (*RYR1*)-related RM and congenital myopathies^62^. Furthermore, both probands exhibited signs of autophagy alterations, including autophagic debris, mitochondrial vacuolization, and myelin figures (**Table 1**, **Figure 1**), which is consistent with the findings of pathophysiological investigations of other RM genetic causes^1,11,46,48,63–65^. Indeed, autophagy insufficiency has emerged as a major cause underlying susceptibility to RM^1,46,48^. In addition, we observed a slight increase in the lipid droplet (LD) content in biopsies from both patients (**Figure 1**, **Table 1**). Although moderate, this finding is consistent with previous reports in patient cells and experimental models of other genetic causes of RM^1,66^, thus underscoring the potential relevance of lipid handling in RM pathology.

**Figure 1:**
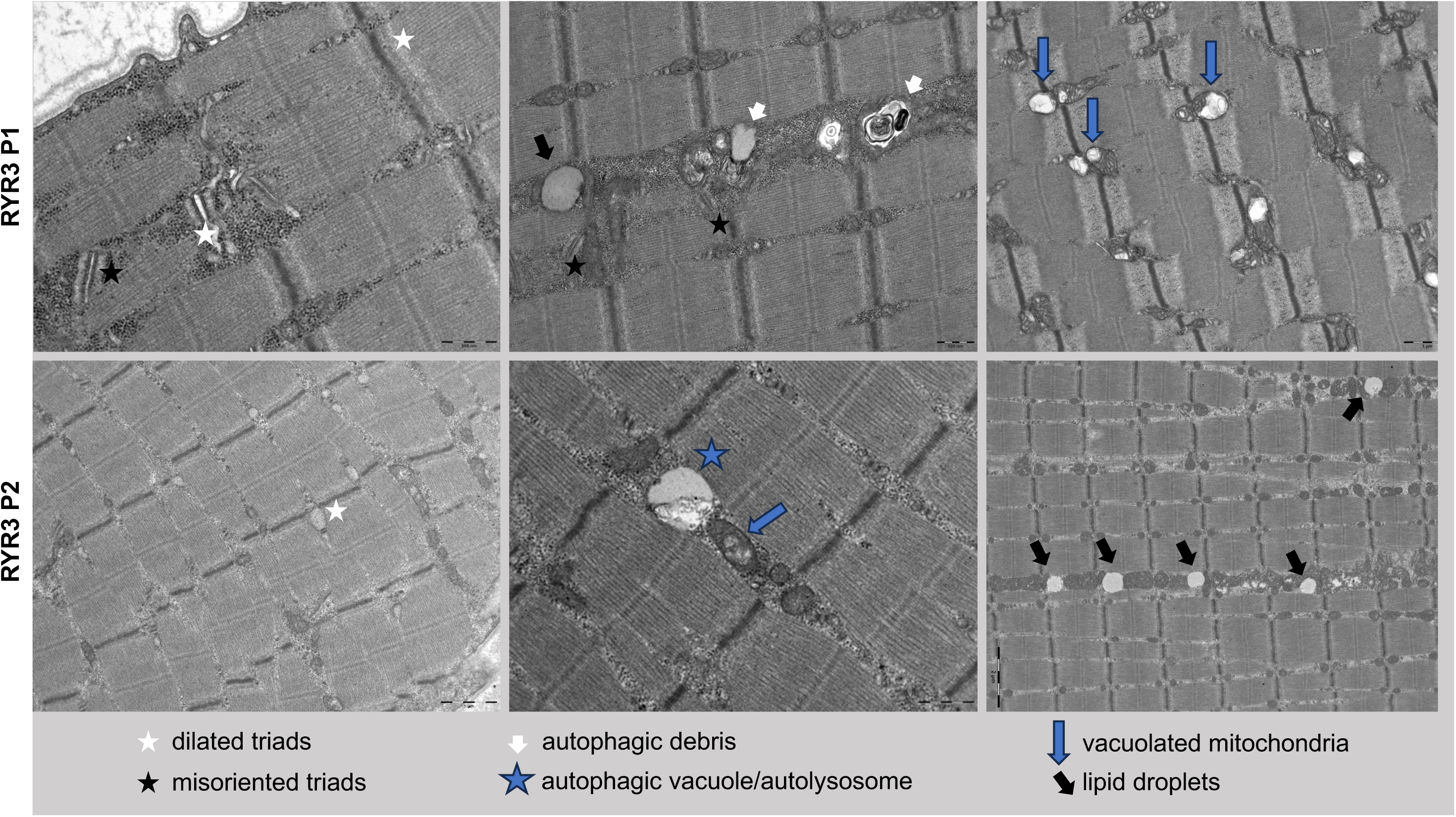
Main findings from ultrastructural investigations. Electron microscopy representative images were obtained from ultrathin sections of deltoid muscle biopsy specimens from *RYR3* P1 and P2. Key features include dilated triads (white star), misoriented triads (black star), autophagic debris (white arrow), autophagic vacuole/autolysosome (blue star), vacuolated mitochondria (blue arrow) and lipid droplets (black arrow).

Overall, both patients exhibited convergent ultrastructural alterations involving ECC structures, autophagy machinery, and mitochondrial integrity. These combined defects indicate a structural and functional vulnerability of skeletal muscle fibers, which is consistent with a pathogenic mechanism underlying susceptibility to RM^67–69^. By analogy with RM associated with *RYR1* variants and other genetic causes of RM, the persistence of triadic disorganization together with evidence of impaired autophagy machinery and mitochondrial network in probands provides mechanistic support for a causal contribution of *RYR3* variants to RM susceptibility.

### *RYR3* variants alter its function

Primary myoblasts from each proband were used to assess the molecular and functional consequences of the identified *RYR3* variants. Quantitative RT-PCR analysis of *RYR3* expression in patients-derived primary myoblasts revealed no significant difference compared with control myoblasts, indicating that the variants do not affect *RYR3* mRNA stability or steady-state transcript levels (**Supplemental Figure 1A**).

To further assess the potential impact of these variants on RyR3 function, we mapped their positions on 3D structures we previously determined for RyR3: the cryo-EM structures full-length mink RyR3 in open and closed forms^59^ and the isolated Repeat34 domain from human RyR3, which was crystallized separately^58^. The mink RyR3 displays more than 96% sequence identity with human RyR3 and the variants and their 3D environments are conserved, thus providing an excellent template for analysis.

For P1, the variants R2669H and N3849H map to the Repeat34 domain and the Central Solenoid (Csol) region, respectively (**Figure 2A**, **B**). The Repeat34 is poorly resolved in full-length cryo-EM reconstructions, but our previous crystal structure of this domain at 1.75 Å resolution (PDB ID: 4ERV) allows for a detailed analysis (**Figure 2C**). Arg2669 is part of a salt bridge network that includes interactions with Glu2662 and Glu2666, which in turn interact with Arg2673. It also forms a hydrogen bond with the side chain of Thr2619, all of which are contained within the same domain. Replacement with a His would thus affect all of these interactions and destabilize this domain. The Repeat34 domain has an enigmatic role in RyR function, but its phosphorylation in RyR1 or RyR2 has been involved in gain-of-function mechanisms and disease^70^, likely by altering the secondary structure content of the domain^71^. Thus, structural perturbations in this domain, induced by the R2669H variant, has the potential to alter function. In contrast, Asn3849 is located in the CSol region. The equivalent residue in mink RyR3 (Asn3837) is located on the surface and interacts with a nearby His side chain in the same domain (**Figure 2D**). As this residue is not located at a domain-domain interaction site, the N3849H may have a minimal functional impact, but it is important to note that a neighboring helix facilitates interactions between the Csol and N-terminal solenoid (NSol). Many disease-causing mutations in RyRs are known to map at domain-domain interactions^3^, so the N3849H could still have a small impact that may be significant in combination with another variant, such as R2669H. In P2, the V610I and V2986I variants map to the N-terminal solenoid (NSol) and Bsol regions, respectively (**Figure 2A**, **B**). Val610 is located at a critical interface between the NSol and Bsol (**Figure 2E**). This interface is allosterically coupled to the pore of the channel. In particular, the equivalent Val610 in mink RyR3 forms van der Waals interactions with Arg2031 in the Bsol (also Arg2031 in human RyR3). Although substitution to Ile is very subtle, adding only a single methyl group can have an important impact on channel gating. For example, in RyR1, the R614C mutation has been involved in malignant hyperthermia^56^ and a cryo-EM analysis has shown that it leads to altered interactions between the Bsol and Nsol regions, resulting in conformational changes that facilitate channel opening^72^. Thus, even a small substitution at this interface may affect the energetics of channel opening. Val2986, equivalent to Val2984 in mink RyR3, is located adjacent to a short flexible loop in the Bsol region (**Figure 2F**). As such, the subtlety of the variant, along with its location, would suggest a minimal impact of this variant on the gating of RyR3. In summary, the R2669H and V610I variants have the highest potential to affect RyR3 function, which can be exacerbated in combination with another variant and thus be involved in the disease phenotype of both patients.

**Figure 2:**
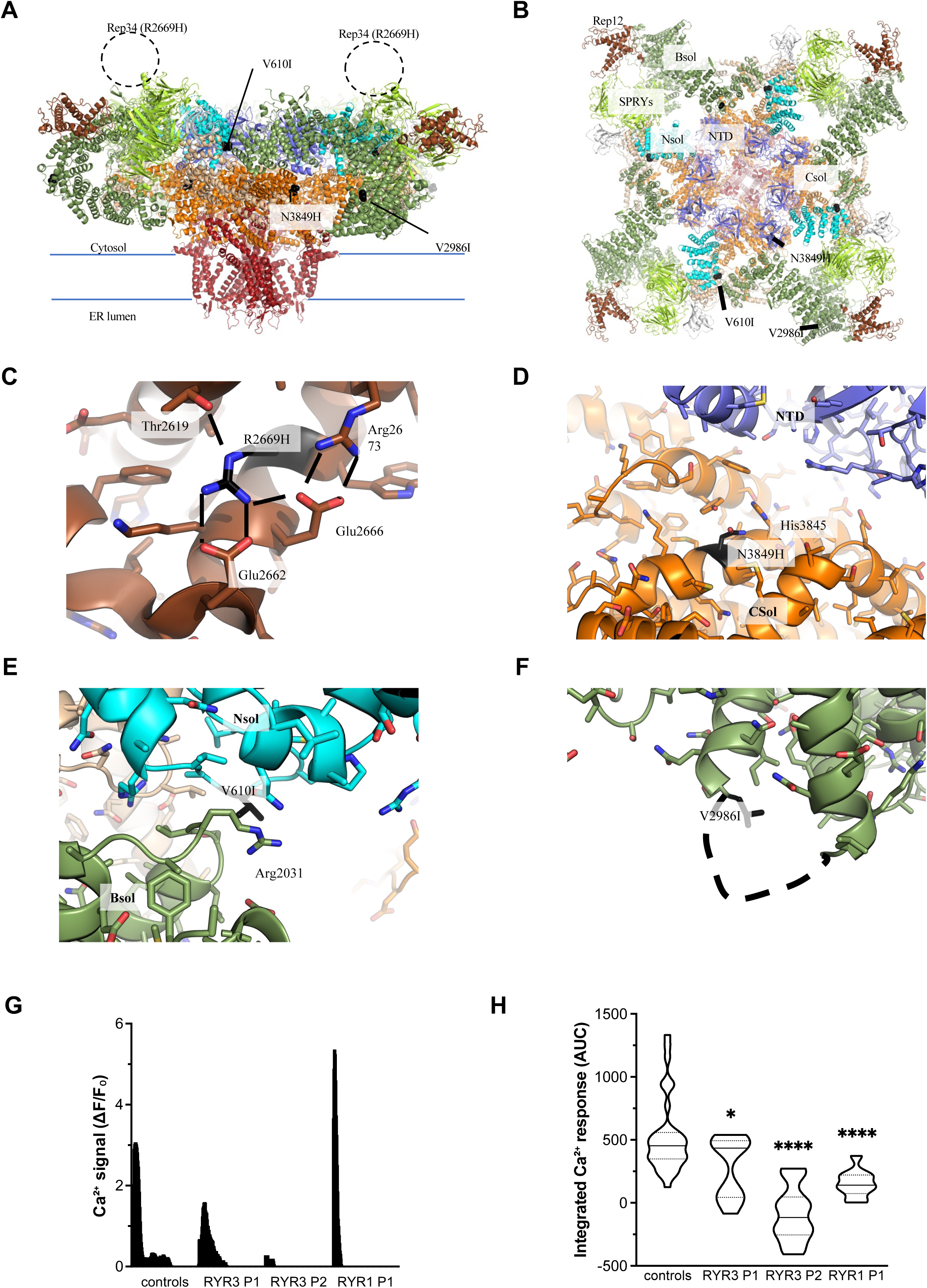
*RYR3* variants alter calcium release **A.** Cryo-EM structure of mink RyR3, as viewed from within the plane of the ER membrane. Different structural regions are colored. Blue: N-terminal domains (NTD); cyan: N-terminal solenoid (NSol); dark green: Bridging Solenoid (BSol); light green: SPRY domains; brown: Repeat12 domain; orange: Central Solenoid (CSol); red: transmembrane region and C-terminal domain. The overall locations of the variants are shown as black spheres and labeled. The dotted circle represents the approximate location of the Repeat34 domain containing the R2669H variant. All residue numbers are for the corresponding human RyR3. **B.** ‘Top’ view of the structure shown in panel A, as seen from the cytosol facing the ER. **C.** Details around the R2669H variant, mapped to the crystal structure of the individual human RyR3 Repeat34 domain. The position of the variant is shown in black. It is involved in a salt bridge network involving Arg2673, Glu2666, and Glu2662, and also forms a hydrogen bond with Thr2619. The salt bridge and hydrogen bonds are represented by dotted lines. Mutation to His will disrupt this network, thus destabilizing the domain. **D.** Close-up around the N3849H variant, located in the Csol (orange). Asn3849 makes interactions with the nearby His3845. Note the interactions between the Csol and the NTD (blue), an interface that is critical for normal RyR function. **E.** Close-up around the V610I variant (black sticks), which is at the interface between the NSol (cyan) and the BSol (green). Val610 interacts with Arg2031. **F.** Details around the V2986I variant (black sticks), located in the BSol domain (green). The dotted line represents a flexible loop immediately adjacent to Val2986. As such, the V2986I variant is less likely to perturb the RyR3 function. **G.** Representative calcium release signal of primary myoblasts from *RYR1* and *RYR3* patients stained with Fluo-4AM and treated with 10 nM 4-chloro-m-cresol (10 minutes recording). The calcium signal is represented as deltaF/F_0_, with F_0_ representing the mean fluorescence level before injection. **H.** Area under the curve (AUC) for the integration of the calcium signal measured from cultured primary myoblasts from *RYR1* and *RYR3* patients. *p < 0.05; ****p < 0.0001 from one-way ANOVA and Dunnett’s post-hoc test.

To confirm the functional impact of *RYR3* variants, we investigated calcium release in cells harboring the *RYR3* variants and compared the responses to control cells and cells from *RYR1*-mutant patient. The cells were stimulated with 4-chloro-m-cresol (4-CmC) to directly activate RyRs channels. Treatment with 10 nM 4-CmC resulted in reduced calcium release in myoblasts derived from both *RYR3* probands and *RYR1* patients (**Figure 1G, H**). Specifically, the peak amplitude of calcium release was significantly decreased in cells from *RYR3* patients (**Figure 1G**), together with a reduced area under the curve (AUC) which was observed in cells from all patients compared to controls (**Figure 1H**).

Overall, these results implicate the identified *RYR3* variants in the RM phenotype of both probands, at least in part by compromising RyR3-dependent calcium signaling in skeletal muscle.

### RyR3 is essential for maintaining skeletal muscle function

RyR3 is expressed during early skeletal muscle development and contributes to the shaping of intracellular calcium signals in differentiating myofibers. While RyR1 is the predominant isoform in adult skeletal muscle, RyR3 is proposed to fine-tune calcium signaling during myogenesis and early postnatal maturation^28,30,73^. Therefore, we first examined the effect of *RYR3* knockdown (KD) on myogenic differentiation in human myoblasts (**Figure 3A, B, Supplementary Figure 1B**). *RYR3* depletion in myoblasts significantly impaired differentiation upon serum withdrawal, as reflected by a marked reduction in the fusion index over a 6-day differentiation period (**Figure 3A, B**).

**Figure 3:**
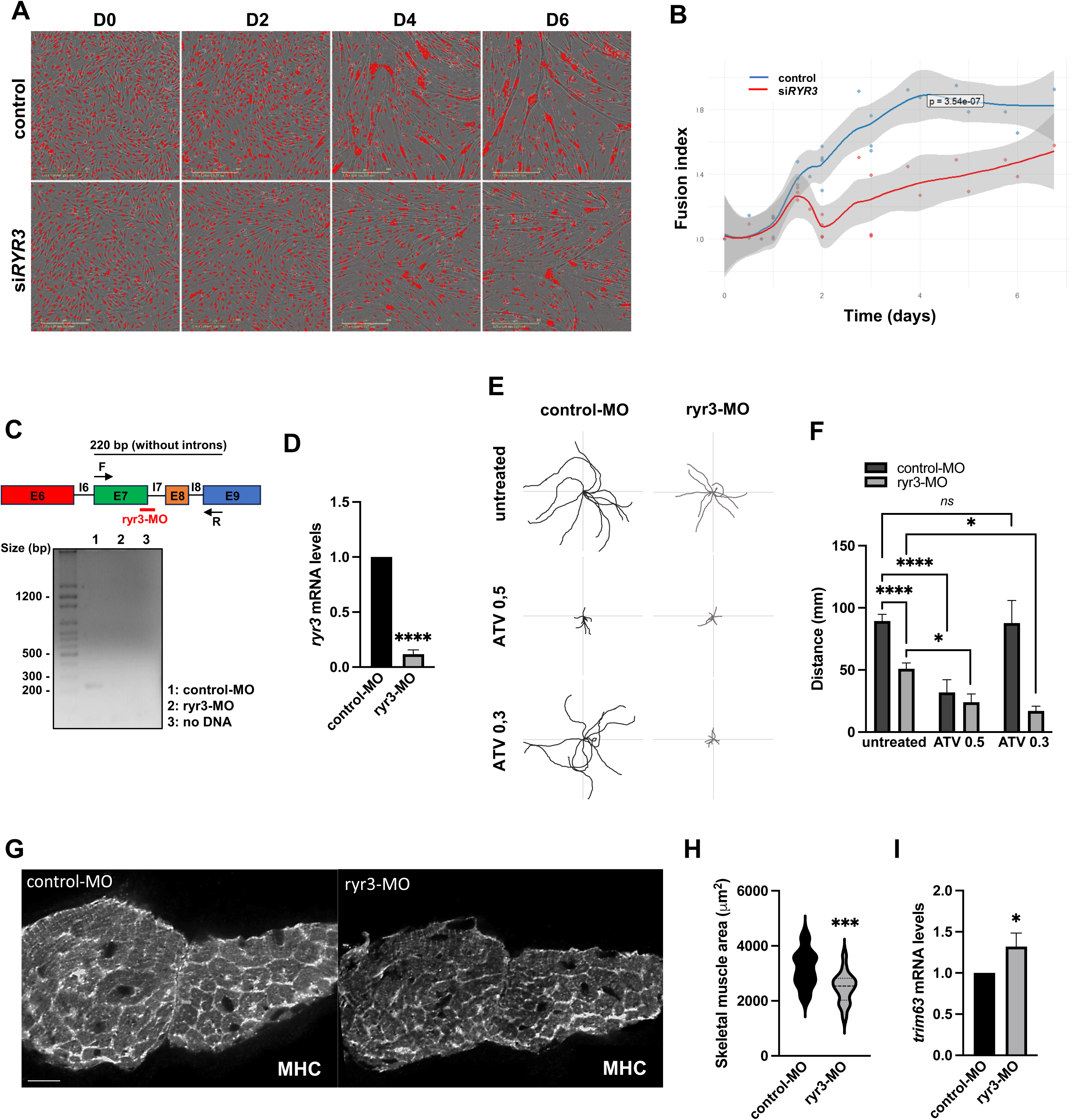
RyR3 is essential for maintaining skeletal muscle function **A.** Differentiation of cultured immortalized myoblasts into myotubes tracked 0 to 6 days post-differentiation and upon transfection of cells with control or *RYR3* siRNA (Nucspot® nuclear stain in red). **B.** Loess regression curve of the myogenic fusion index measuring the ability of control or si*RYR3* transfected myoblasts to form multinucleated myotubes. **C.** Design of the antisense MO altering mRNA splicing targeting the splicing region (exon 7-intron 7) in *ryr3* transcripts. The flanking primers amplifying a 220 bp cDNA region are indicated with black arrows. Agarose gel electrophoresis of RT-PCR products from cDNA of control-MO and ryr3-MO injected zebrafish embryos illustrating mRNA decay following splicing defect. **D.** RT-qPCR measurement of *ryr3* expression in control-MO and ryr3-MO injected zebrafish embryos. ****p < 0.0001 from Student’s t test. **E.** Representative traces of zebrafish larval motor behavior with a touch-evoked escape response (TEER). 48 hours post-fertilization (hpf) larvae injected with control-MO or ryr3-MO were treated with 0.3 μM or 0.5 μM atorvastatin (ATV) in zebrafish larval water for 24h prior testing. **F.** Swimming distance measurement of TEER from control-MO or ryr3-MO injected larvae treated with 0.3 μM or 0.5 μM ATV or non-treated controls. Data are represented as mean + SEM, *p < 0.05; ****p < 0.0001 from two-way ANOVA followed by Tukey’s post hoc test. **G.** Immunofluorescence labeling of myosin heavy chain (MHC) in myotome cross-sections from 48 hpf zebrafish larvae injected with control-MO or ryr3-MO. Scale bar represents 20 µM. **H.** Skeletal muscle area measurements of MHC-stained myotome from 48 hpf control-MO or ryr3-MO-injected larvae. ***p < 0.001 from Student’s t test. **I.** RT-qPCR measurement of *trim63*(MuRF1) expression in control-MO and ryr3-MO injected zebrafish larvae. Data are presented as mean SD, *p < 0.05 from Student’s t test.

To further investigate the role of Ryr3 in skeletal muscle integrity and function, we developed a zebrafish model. In zebrafish, five genes encode ryanodine receptor (RyR) isoforms: *ryr1a*, *ryr1b*, *ryr2a*, *ryr2b* and *ryr3*. *ryr1a*, *ryr1b* and *ryr3*, are expressed in the myotome of the zebrafish embryos^74^. Zebrafish Ryr3 displays strong evolutionary conservation with human RyR3, exhibiting 77% amino acid sequence identity. Previous investigations in zebrafish have shown that loss of *ryr3* modulates contractile activity and skeletal muscle fiber maturation^29^. Here, we used a morpholino targeting a splicing region conserved across all four transcripts of *ryr3* (exon 7-intron 7 of the XM_009294775.5 canonical transcript) to inhibit its expression (**Figure 3C**). Skipping exon 7 is predicted to cause a frameshift downstream, likely introducing a premature termination codon and triggering nonsense-mediated decay of the transcript. Gel electrophoresis of the targeted amplicon revealed loss of the expected band in the *ryr3*-MO condition, supporting the efficacy of *ryr3* inhibition (**Figure 3C**), which was further confirmed by RT-qPCR (**Figure 3D**). By 48 hpf, zebrafish muscles are fully differentiated, with established myotomes, and larvae display stereotyped escape responses to touch, enabling assessment of muscle performance via the Touch-Evoked Escape Response (TEER) test^75^.

In zebrafish, the RM phenotype is characterized by reduced locomotor activity and altered muscle morphology^48,76,77^. Then, we evaluated locomotor activity in 50 hpf larvae by recording individual swimming episodes in ryr3-MO zebrafish larvae. As a result, we found that *ryr3* KD larvae exhibited reduced trajectories and swimming distance compared to controls (**Figure 3E, F**). To validate the specificity of this phenotype to RM pathology, 30 hpf larvae were exposed to atorvastatin (ATV), a known pharmacological RM inducer previously shown to elicit RM-related features in zebrafish^48,76^. TEER quantification revealed that ATV exacerbated locomotor defects in ryr3-MO larvae in a dose-dependent manner. At the highest dose, both control and *ryr3* KD larvae exhibited reduced swimming distance, whereas at the lower dose, motor deficits were observed only in ryr3-MO larvae. In addition, immunofluorescence analysis of transverse cryosections from 50 hpf zebrafish via an anti-myosin heavy chain (MHC) antibody revealed that *ryr3* morphants presented disorganized skeletal muscle fibers and a significantly reduced myotome area compared to controls (**Figures 3G, H**). Consistent with this morphological alteration, expression of the muscle-specific atrophy-related gene *murf1* (*trim63*) was significantly increased by 32% (**Figure 3I**), reflecting early or mild signs of muscle atrophy. Finally, Oil Red O (ORO) staining of transverse cryosections from 50 hpf zebrafish revealed that ryr3 morphants display an accumulation of lipid droplets within skeletal muscle (**Supplementary Figure 2A, B**). This observation is consistent with our ultrastructural analyses (**Figure 1**) and ORO staining (**Supplemental Figure 2C**) performed on muscle samples from *RYR3* patients, as well as with previous reports describing lipid accumulation in other genetic contexts associated with RM predisposition^66,67^.

Collectively, these findings demonstrate that RyR3 is essential for skeletal muscle development and function. Loss of *RYR3* impairs skeletal muscle fiber formation *in vitro* and disrupts muscle function and structural integrity *in vivo*. Together, these results provide strong functional evidence linking RyR3 deficiency to muscle pathology and further support its role in RM susceptibility.

### Autophagy defects underlie RYR3-related RM

To assess cellular vulnerability to metabolic stress, primary myoblasts from patients and controls were subjected to 4h of nutrient deprivation. As shown in **Figure 4A**, control cells exhibited limited cell death under starvation conditions. In contrast, patient-derived myoblasts showed a significantly higher level of cell death when exposed to the same treatment. These findings indicate that patient-derived myoblasts display increased sensitivity to nutrient deprivation, which is consistent with a defective cellular response to metabolic stress.

**Figure 4:**
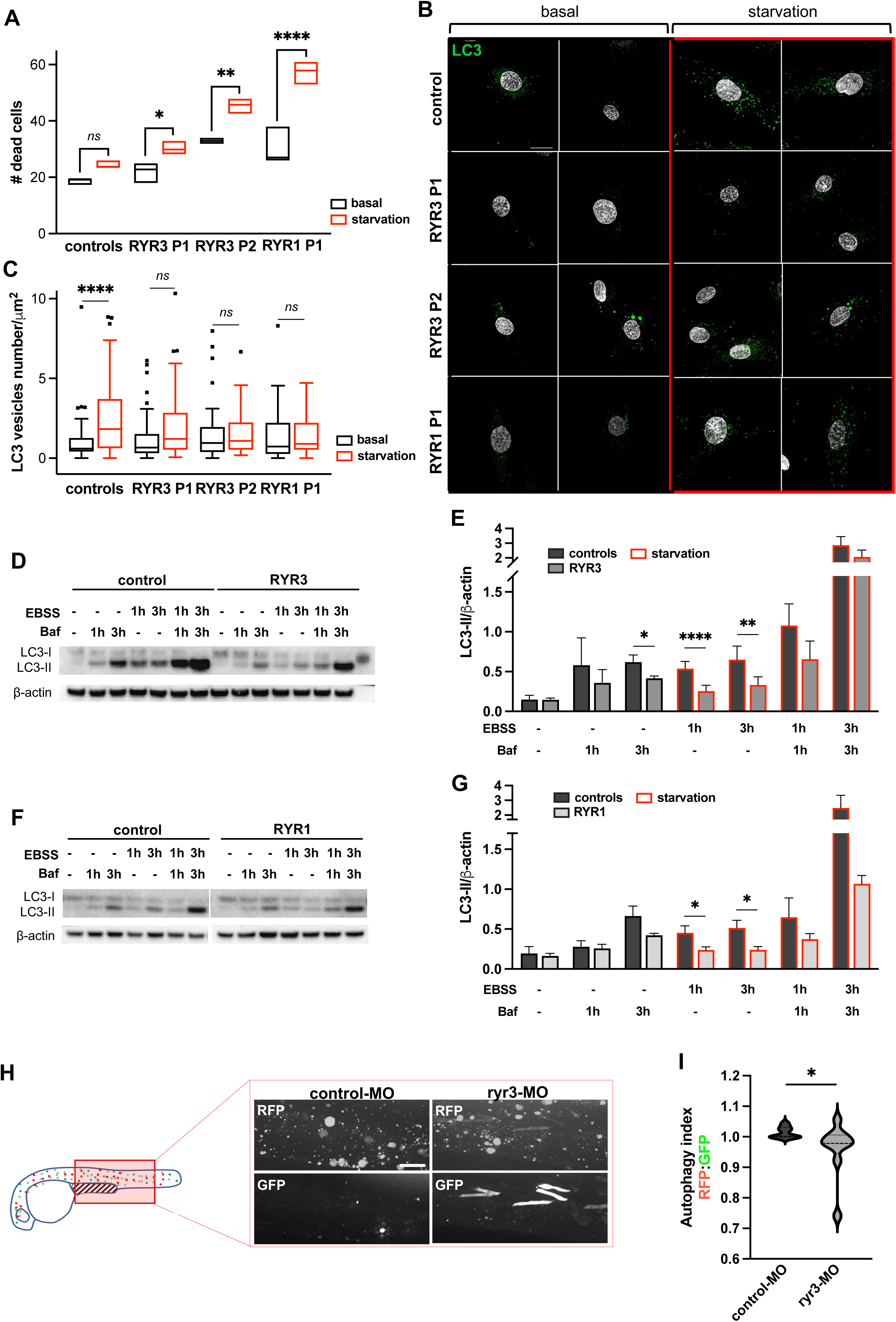
RyR3 deficiency induces autophagy defects **A.** Quantification of control and patients primary myoblasts cell death under basal culture conditions and 4h starvation (EBSS). *p < 0.05, ***p < 0.001, ****p < 0.0001, *ns*, non-significant from 2-way ANOVA and Sidak’s post hoc test. **B-C.** Representative images (**B**) and quantification (**C**) of LC3 positive vesicles in control and patients’ primary myoblasts cultured in basal or 2h starvation conditions. ****p < 0.0001, *ns*, non-significan from two-way ANOVA followed by Sidak’s post hoc test. **D-E.** Representative western blotting (**D**) and quantification (**E**) of LC3-II in control and *RYR3* patients’ primary myoblasts starved (EBSS) or incubated with the autophagosome-lysosome fusion inhibitor bafilomycin A1 (Baf) for indicated time points. Data are represented as mean + SD, *p < 0.05, **p < 0.01, ****p < 0.0001 from two-way ANOVA and Bonferonni post-hoc test. **F-G.** Representative western blotting (**F**) and quantification (**G**) of LC3-II in control and *RYR1* patients primary myoblasts starved (EBSS) or incubated with bafilomycin A1 (Baf) for indicated time points. Data are represented as mean + SD *p < 0.05, **p < 0.01, ****p < 0.0001 from two-way ANOVA and Bonferonni post-hoc test. **H-I.** Confocal imaging of skeletal muscle (**H**) and autophagy index quantification of the normalized RFP to GFP signal (**I**) of larvae co-injected with the autophagic GFP-LC3-RFP-LC3ΔG probe and control-MO or ryr3-MO. *p < 0.05 from Student’s t-test.

Skeletal muscle copes with metabolic stress by activating autophagy^78–80^. Defects in autophagy have been implicated in a broad spectrum of muscle diseases, including muscle atrophy and myopathies, and have also been directly associated with increased susceptibility to RM in genetic disorders such as *LPIN1* and *TANGO2* deficiencies, highlighting the critical role of this pathway in protecting muscle fibers against metabolic and energetic stress^1,46–48^.

Autophagosome biogenesis and autophagic flux rely on the integrity of endoplasmic/sarcoplasmic reticulum membranes and on tightly regulated intracellular calcium signaling^49,50,81^. In light of the structural abnormalities of the SR observed in skeletal muscle biopsies from patients (**Figure 1A**) and the calcium dysregulation detected in *RYR3*-deficient cells (**Figure 2G**, **H**), we examined autophagy dynamics under starvation conditions.

Autophagosome formation was assessed by immunofluorescence detection of LC3 puncta under basal conditions or after 2h of starvation. In control myoblasts, nutrient deprivation induced robust accumulation of autophagosome LC3-positive vesicles, which was consistent with the activation of autophagy (**Figure 4B, C**). In contrast, patient cells presented no significant difference in the number of autophagosomes between basal and stress conditions, highlighting the defective adaptation of myoblasts to starvation (**Figure 4B, C**). To further assess autophagic function, we evaluated LC3 lipidation by immunoblotting at multiple starvation time points in the presence or absence of bafilomycin A1 (**Figure 4D, E**). In control cells, starvation induced a progressive increase in the LC3-II level, which was further augmented upon bafilomycin treatment, indicating ongoing autophagosome formation and lysosomal degradation (**Figure 4D, E**). In contrast, *RYR3* P1 myoblasts displayed a markedly attenuated accumulation of LC3-II over time, both under starvation alone and in the presence of bafilomycin, which was consistent with impaired autophagic flux (**Figure 4D, E**). A similar reduction in LC3-II accumulation was observed in P2 (**Supplementary Figure 3A**) and in *RYR1*-deficient myoblasts (**Figure 4F, G**), supporting a reproducible defect in starvation-induced autophagy across models. To further validate these findings, LC3 levels were measured following siRNA-mediated KD of *RYR3* in control myoblasts subjected to starvation. Compared with siRNA control, *RYR3* silencing resulted in reduced LC3 levels (**Supplementary Figure 3B**), confirming that *RYR3* depletion is sufficient to impair starvation-induced autophagic responses. Taken together, these findings support a role for RyR3 in the autophagic activation during metabolic stress in skeletal muscle cells.

To determine whether autophagy impairment is conserved upon *ryr3* inhibition in zebrafish, we assessed autophagic flux using the GFP-LC3-RFP-LC3ΔG fluorescent reporter^61^. Upon autophagy induction, the probe is cleaved into GFP-LC3, which is incorporated into autophagosomes and degraded in lysosomes, and cytosolic RFP-LC3ΔG, which serves as an internal control. The autophagy index was quantified from confocal images acquired in skeletal muscle by calculating the RFP/GFP fluorescence ratio, reflecting the autophagic flux efficiency (**Figure 3H**, **I**). We observed a significant decrease in autophagy efficiency in ryr3-MO zebrafish larvae compared with controls (**Figure 3H**, **I**), a phenotype similar to that reported in the *tango2* KD model^48^, further suggesting that *ryr3* loss of function recapitulates the mechanistic defects associated with *RYR3* variants.

Overall, our data indicate that RyR3 acts as a key modulator of starvation-induced autophagic responses in skeletal muscle. These findings provide functional evidence that RyR3 disruption predisposes to RM and further supoprt the contribution of autophagy impairment to RM disease pathophysiology.

### RyR3 deficiency is associated with mitochondrial dysfunction

Defects in autophagy may compromise mitochondrial quality control in skeletal muscle, as efficient removal of damaged organelles is required to preserve bioenergetic capacity and metabolic flexibility. Impaired autophagic flux can lead to the accumulation of dysfunctional mitochondria, increased oxidative stress, and defective metabolic adaptation—mechanisms implicated in inherited myopathies and genetic forms of RM^44,45,79^. Consistent with this framework, ultrastructural analysis of patient muscle biopsies in our study supports incomplete mitophagy (**Figure 1A**, **Table 1**), which may alter mitochondrial quality control and compromise mitochondrial function.

Beyond autophagic regulation, mitochondrial function is influenced by SR-mitochondria contact sites that enable localized calcium transfer from the SR to adjacent mitochondria, thereby enhancing oxidative phosphorylation during periods of increased energetic demand^51,52^. Importantly, RyR1 and RyR2 have been shown to functionally participate in calcium transfer within these microdomains^51,53–55^. We therefore examined whether RyR3 deficiency similarly impacts mitochondrial function. Measurement of oxygen consumption rate (OCR) in primary myoblasts revealed reduced respiration in *RYR3* and *RYR1* cells compared with controls (**Figure 5A, Supplementary Figure 4 A-C**), indicating impaired mitochondrial oxidative capacity. To further evaluate mitochondrial function, mitochondrial membrane potential was assessed using tetramethylrhodamine ethyl ester (TMRE) staining (**Figure 5B**). Under basal conditions, the mitochondrial membrane potential was not significantly reduced in *RYR3* patient-derived myoblasts when compared to controls. Notably, nutrient deprivation further exacerbated this decrease, indicating that RyR3-deficient mitochondria fail to maintain membrane polarization under this stress. Following FCCP treatment, which collapses the proton gradient, RyR3-deficient cells presented a lower residual TMRE signal, which was consistent with an overall reduction in mitochondrial membrane potential. Together, these findings support impaired mitochondrial function and a reduced capacity to adapt to energetic stress in RyR3-deficient myoblasts.

**Figure 5:**
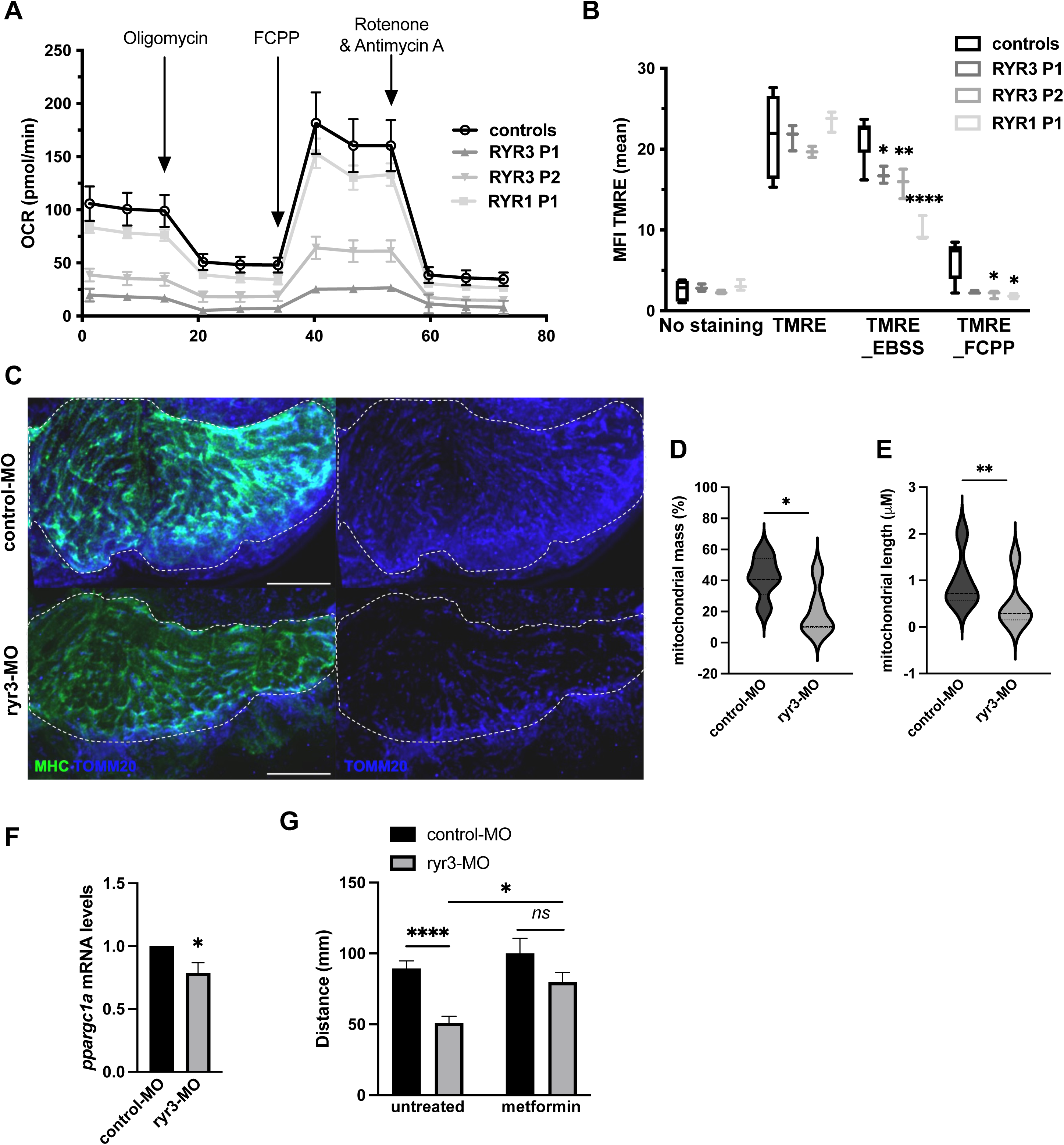
RyR3 deficiency causes mitochondrial dysfunction **A.** Oxygen consumption rate (OCR) measurement in control and patients’ primary myoblasts upon stimulation with oligomycin, carbonyl cyanide-p-trifluoromethoxyphenylhydrazone (FCCP) and rotenone and antimycin. **B.** Fluorescent quantification of mitochondrial transmembrane potential using tetramethylrhodamine ethyl ester (TMRE) staining in control and patients’ primary myoblasts starved (EBSS) or treated with FCCP. *p < 0.05, ****p < 0.0001, *ns*, non-significant from two-way ANOVA and Dunnett’s post hoc test. **C.** Mitochondria staining by immunofluorescent labeling of myosin heavy chain (MHC, green) and TOMM20 (blue) in myotome cross-sections from 48 hpf control-MO or ryr3-MO injected zebrafish. Scale bar represents 20 µM. **D-E.** Mitochondrial mass (**D**) and length (**E**). *p < 0.05 and **p < 0.01 from Student’s t-test. **F.** RT-qPCR quantification of *ppargc1a* expression in control-MO and ryr3-MO injected zebrafish embryos. Data are presented as mean +SD and *p < 0,05 from Student’s t-test. **G.** Swimming distance measurements of TEER from control-MO or ryr3-MO injected larvae treated with 50 μM metformin for 24 hours or non-treated controls. Data are presented as mean +SEM *p < 0.05; ****p < 0.0001 from two-way ANOVA and Tukey’s post-hoc test.

We next examined mitochondrial organization in *ryr3*-deficient zebrafish. Immunofluorescence staining of TOMM20 revealed an abnormal mitochondrial network within the myotome of the *ryr3* morphants (**Figure 5C**). Quantitative analysis showed a reduction in mitochondrial mass (**Figure 5D**) together with decreased mitochondrial length (**Figure 5E**). These structural abnormalities are consistent with the bioenergetic defects observed in *RYR3* patient-derived myoblasts, supporting impaired mitochondrial homeostasis across models.

PGC1-α is a master regulator that coordinates transcriptional programs controlling mitochondrial mass, oxidative metabolism, and respiratory function in skeletal muscle^82,83^ Given that calcium-dependent signaling pathways regulate PGC1α/*PPARGC1A* expression in skeletal muscle^84–86^, we hypothesized that *ryr3* deficiency might alter its expression. Accordingly, we found that *ryr3* morphants presented reduced *ppargc1a* transcript levels (**Figure 5F**), which is consistent with impaired engagement of mitochondrial biogenesis pathways and in line with the reduced mitochondrial mass and bioenergetic defects observed across models.

Given the central role of AMP-activated protein kinase (AMPK) signaling in coordinating energy sensing, mitochondrial biogenesis, and autophagy, we next evaluated whether pharmacological activation of this pathway could ameliorate the muscle phenotype^87–89^. Treatment of *ryr3*-deficient larvae with metformin, an AMPK activator and upstream regulator of PGC1α, improved the locomotor performance of ryr3-MO zebrafish larvae (**Figure 5G**).

Together, these results indicate that RyR3 deficiency disrupts mitochondrial bioenergetics and organelle homeostasis in skeletal muscle, suggesting a link between SR calcium flux dysregulation and defective autophagy, as well as altered metabolic adaptation.

## Discussion

Although more than 1200 *RYR3* variants and polymorphisms have been reported in public databases (some associated with epileptic encephalopathy), their pathogenic relevance has thus far been convincingly established only in a limited number of cases, notably in congenital myopathies and arthrogryposis^26,27^. Here, we showed that patients carrying the recessive variants Arg2669His and Asn3849Hisp (P1) and Val610Ile and Val2986Ile (P2) presented convergent alterations involving ECC, autophagy and the mitochondrial compartment, leading to a functional vulnerability of muscle fibers.

Although RyR3 is not the principal excitation-contraction coupling (ECC) channel in adult skeletal muscle, the triadic disorganization observed in patients with *RYR3* variants parallels the structural alterations described in *RYR1*-related RM and congenital myopathies, suggesting that RyR3 deficiency may destabilize triadic architecture, reinforcing its causal role in RM susceptibility and strongly supporting the inclusion of these cases within the spectrum of triadopathies^62^. Our structural mapping of the variants supports an oligogenic-like contribution of two *RYR3* variants per patient which is reminiscent of modifying alleles reported in *RYR1*-related disorders, where variants combinations can modulate disease severity or stress susceptibility^18,90,91^.

The comparable reduction in 4-CmC-induced calcium release observed in myoblasts from both *RYR3* patients supports a functional perturbation of RyR3 and may reflect altered channel gating, as previously described for the *RYR1* variant studied here^92^. Previous experimental *RYR3* ablation studies have shown that RyR3 deficiency reduces the calcium spark frequency and modulates intracellular calcium dynamics^28,93^. Importantly, *RYR1* mutations can either increase or reduce calcium release and both gain and loss of function mechanisms have been associated with muscle diseases^15,19,91,94^.

*RYR3* depletion impaired myoblast fusion *in vitro* and induced an atrophy-like phenotype in zebrafish, characterized by reduced locomotor performance, decreased myotome size, and increased expression of the atrogene MurF1 (*trim63*). Furthermore, zebrafish data revealed that *ryr3*-deficient larvae exhibit increased sensitivity to ATV-induced muscle stress, suggesting that functional RyR3 is required for stress resilience.

Given the recently reported activating effect of ATV on RyR1, the increased sensitivity observed in ryr3-deficient larvae may reflect an imbalance in RyR-mediated calcium signaling, potentially influencing calcium dynamics mediated by RyR1 zebrafish orthologues (ryr1a/ryr1b) in the absence of ryr3^95^.

Skeletal muscle relies heavily on autophagy to maintain energy balance and to remove damaged components during metabolic stress. By contrast, autophagy insufficiency has emerged as a key mechanism underlying RM susceptibility^1,46,48^. Interestingly, RyR1 dysfunction has been linked to altered autophagic flux and mitochondrial abnormalities in congenital myopathies^96–98^. Consistent with this framework, we found here that both RyR3- and RyR1-deficient myoblasts failed to properly induce autophagosome formation upon starvation and exhibited impaired autophagic flux. These findings indicate that RyR-mediated calcium release is required for efficient activation of autophagy under metabolic stress conditions. In the context of RyR3 deficiency, abnormal SR calcium signaling likely compromises starvation-induced autophagic responses, thereby increasing cellular susceptibility to metabolic challenge. The similar impairment observed in *RYR1*-deficient cells further supports a broader role for RyR-dependent calcium signaling in autophagic activation and positions *RYR3* within the growing group of RM-associated genes regulating autophagy-mediated stress adaptation pathways.

Mitochondrial integrity is critical for skeletal muscle performance and adaptation. Impaired mitophagy or mitochondrial bioenergetics contributes to inherited myopathies and metabolic stress disorders^99^. Consistent with defective autophagic quality control, patient biopsies presented features compatible with incomplete mitophagy. Functionally, RyR3-deficient myoblasts displayed reduced basal and maximal oxygen consumption and decreased mitochondrial membrane potential, particularly under nutrient deprivation. In zebrafish, *ryr3* loss resulted in reduced mitochondrial mass, and decreased *ppargc1a* expression, indicating compromised mitochondrial biogenesis. Beyond quality control, SR-mitochondria contact sites enable localized calcium transfer that stimulates oxidative phosphorylation during increased energetic demand^51,52^. Although a role for RyR3 at contact sites remains to be demonstrated, this study suggests that impaired calcium-dependent organelle crosstalk underlies metabolic vulnerability in RyR3 deficiency.

Overall, our study establishes *RYR3* as a novel genetic contributor to recurrent RM and highlights its central role in skeletal muscle stress adaptation. By integrating human genetics, patient cell analyses, and zebrafish models, we demonstrate that RyR3 deficiency disrupts autophagy, impairs mitochondrial homeostasis, and destabilizes triadic architecture, collectively reducing muscle resilience to metabolic stress. These findings not only expand the spectrum of RyR-related muscle disorders but also underscore a broader mechanistic principle: efficient SR calcium signaling is essential for coordinating organelle crosstalk and adaptive responses in skeletal muscle. Ultimately, this work positions RyR3 within the growing group of RM-associated genes regulating calcium-mediated autophagic and mitochondrial pathways, providing a framework for understanding disease susceptibility and potential therapeutic targeting.

## Data Availability

All data produced in the present study are available upon reasonable request to the authors

## Acknowledgments

The authors thank Meriem Garfa from the cell-imaging platform for technical assistance in confocal imaging acquisition and analysis, Ivan Nemazzany from the metabolic platform SFR-Necker for assisting with the SeaHorse experiments, the MyoLine platform for immortalizing of human cells, Christine Bole-Feysot for exome analysis, Caroline Tuchmann-Durand for regulatory aspects and Norma Romero for her contribution to the histopathological analyses. This work was supported by grants from Agence Nationale de la Recherche (ANR-AAPG 2018 CE17 MetabInf, ANR-AAPG CE17 2023 Rhabdophagy), the Association Française contre les Myopathies (AFM 19773, AFM 24269, AFM 29118), and patient associations (Nos Anges, AMMI, OPPH, No Myolyse, Des ailes pour L, Hyperinsulinisme, Noa Luû).

---

**Supplementary Figure 1.**
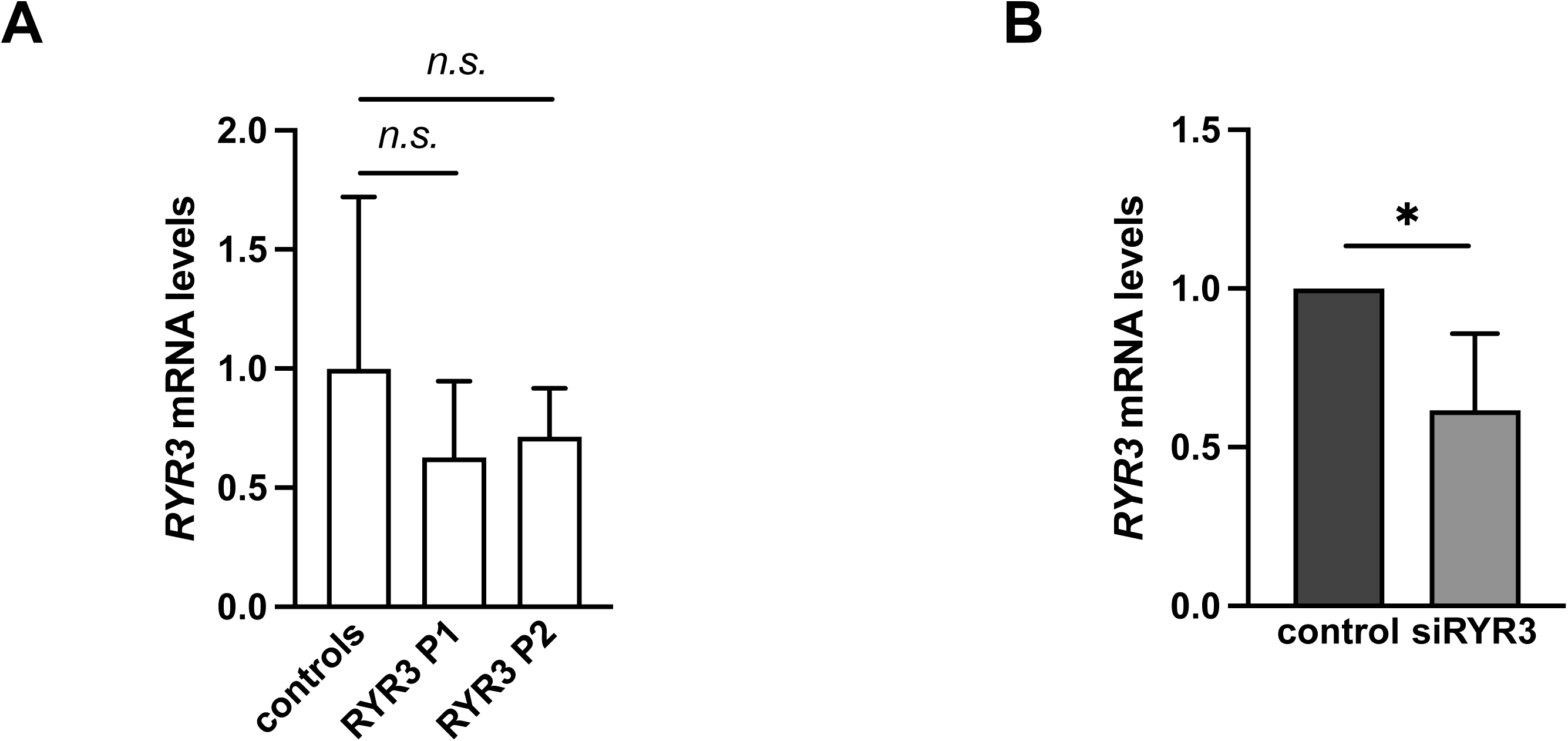
**A.** Relative *RYR3* mRNA levels in controls, *RYR3* P1 and *RYR3* P2 primary myoblasts. **B.** Relative *RYR3* mRNA levels in control primary myoblasts 72 hours after transfection with siRNA targeting *RYR3* expression. Data are plotted as mean + standard deviation. n.s. non significant; *p<0,05 from paired t-tests.

**Supplementary Figure 2.**
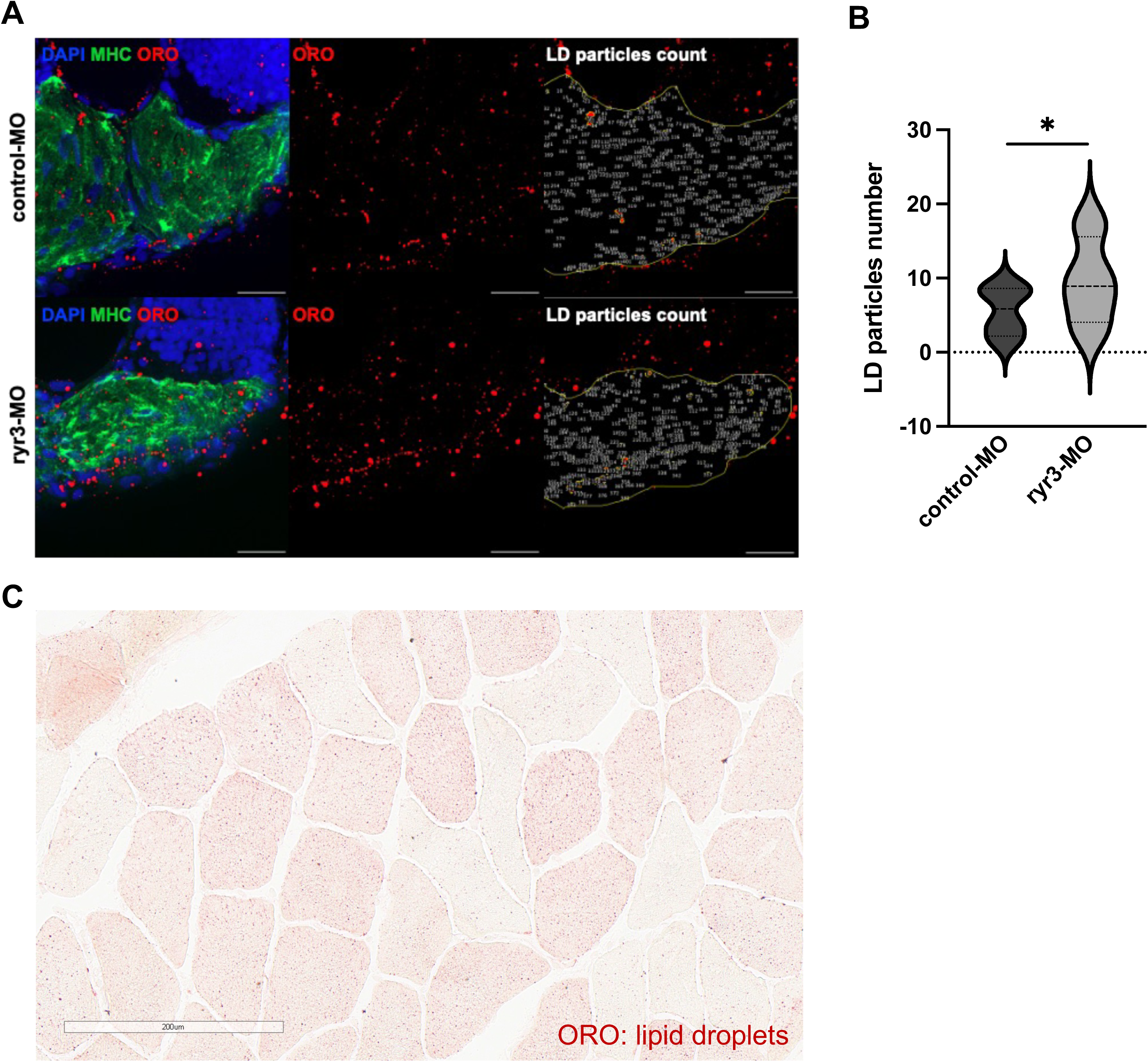
**A.** Representative transversal cryosections of skeletal myotomes from 50 hours post fertilization zebrafish larvae stained for myosin heavy chain (MHC, skeletal muscle) and Oil Red O (ORO, neutral lipids). Right panel shows indiviual detected lipid droplets (LD) particles within the outlined skeletal muscle. Scale bar represents 20 µM. **B.** Violin plot of lipid droplets particles number. *p<0,05 from paired t-test. **C.** Skeletal muscle biopsy from *RYR3* P2 stained with ORO showing neutral lipid content at the upper limit of normal.

**Supplementary Figure 3.**
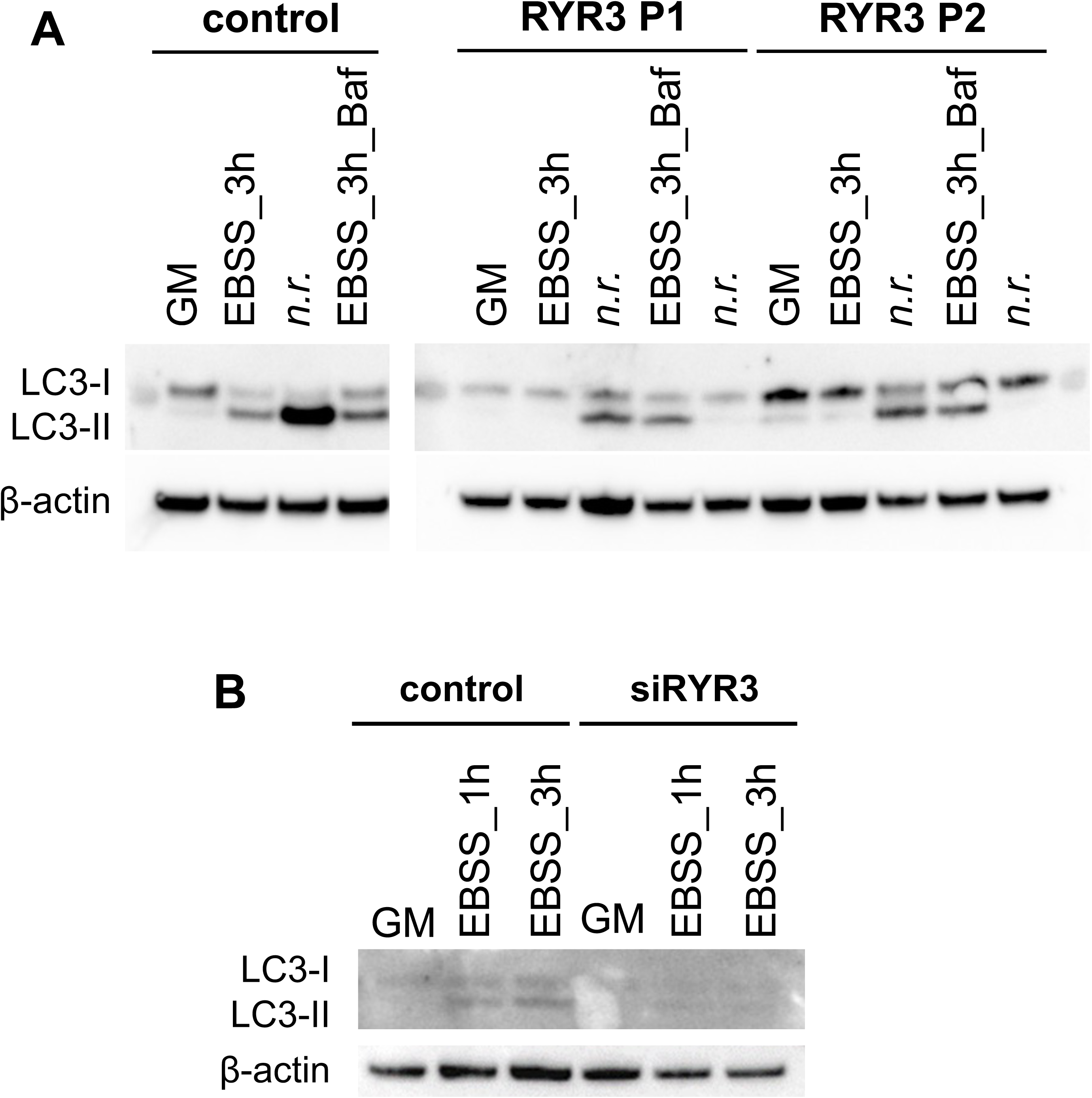
**A.** Representative immunoblot of LC3 lipidation upon starvation (EBSS: Earle’s Balanced Salt Solution) and Bafilomycin A1 treatment (Baf) in primary myoblasts from *RYR3* P1 and *RYR3* P2 patients. Due to limited proliferative capacity of primary cells, the experiment could only be repeated twice. GM: Growth Medium. β-actin was used as a loading control. n.r. (non relevant) indicates lanes that were included on the blot but are not relevant for the present study. **B.** Representative immunoblot of LC3 lipidation upon starvation in siRNA-mediated *RYR3* depleted primary myoblasts. β-actin was used as a loading control.

**Supplementary Figure 4.**
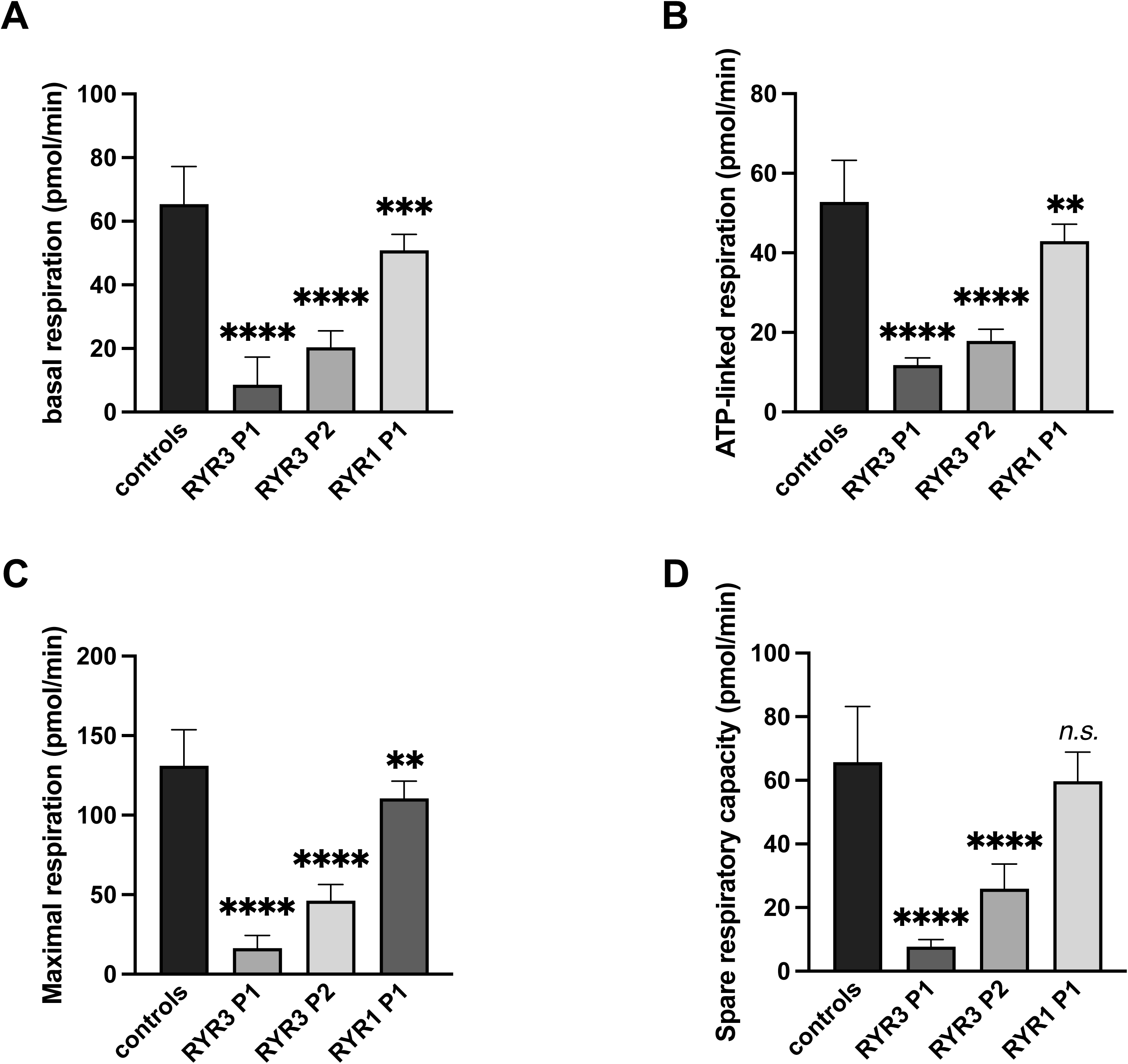
**A.** Basal respiration calculated as the OCR prior to oligomycin injection minus non-mitochondrial respiration. **B.** ATP-linked respiration, determined as the decrease in OCR following oligomycin injection. **C.** Maximal respiration, calculated as the OCR following FCCP injection minus non-mitochondrial respiration. **D.** Spare respiratory capacity, determined as maximal respiration minus basal respiration. Data are presented as mean + SD from n = 3 independent experiments. *n.s.* non significant, **p<0.01 and **** p<0.0001 from one-way ANOVA followed by Dunnett’s multiple comparisons.

